# Predicting the future SARS-COV-2 reproductive rate

**DOI:** 10.1101/2020.08.09.20170845

**Authors:** DSJ Ellis, DI Papadopoulos, I Donkov, S Bishara

## Abstract

**Aims:** To generate a predictive model for the SARS-COV-2 viral reproductive rate, based on government policy and weather parameters.

**Methods:** A multivariate model for the log_10_ of viral reproductive rate was constructed for each country using lockdown stringency (Oxford University tracker), temperature and humidity, for the 1^st^ 110 days of 2020. This was validated by extrapolating to the following 51 days, and comparing the predicted viral rate and cumulative mortality with WHO data.

The country models was extrapolated to July 2021 using projected weather forecast for four scenarios; continuing with the 11/6/2020 lockdown policy, 100% lockdown, 20% lockdown and no lockdown.

**Results:** From pooled data (40 countries), lockdown stringency had a strong negative correlation with log_10_viral reproductive rate (−0.648 at 21 days later). Maximum temperature correlated at -0.14, 14 days later and humidity correlated at +0.25, 22 days later. Predictive Models were generated for 11 countries using multivariate regression of these parameters. The R^2^ correlation for log_10_R_0_ ranged from 0.817 to 0.987 for the model generation period. For the validation period, the Pearson’s coefficient of correlation ranged from 0.344 to 0.984 for log_10_R_0_ and from 0.980 to 1.000 for cumulative mortality.

Forward extrapolation of these models for 5 nations, demonstrate, that removing the lockdown will result in rapid spread of the disease ranging from as soon as July 2020 for Russia, UK, Italy and India to January 2021 for the USA. The current (11/6/20) lockdown in the USA, Spain, UK, France, Germany, Turkey can control the disease but other nations will need to intensify their lockdowns to prevent future resurgence. Most nations will require more stringent lockdowns in January than in July.

**Conclusion:** The viral reproductive rate is highly predicted by a combination of lockdown stringency, temperature and humidity. Country specific predictive models can provide useful forecast of policy requirements.

## Introduction

The Sars-CoV-2 outbreak started initially in Wuhan and spread rapidly around the world. It was declared a global pandemic by the World Health Organisation in March 2020. As of the 12^th^ of July 2020 there have been over 12,000,000 confirmed cases and 500,000 deaths, [1] though the true number of cases is likely to be much higher due to the current limitations of testing methods. A better indication of the true infection rate can be gleaned from antibody prevalence studies in national sample populations, which may place the cumulative incidence of infection at greater than 10 times higher than the confirmed infection rate,[2] typically associated with an infection fatality rate of between 0.5% and 1%.[3]

As there are no highly effective treatments or a vaccine, the national response to the pandemic has centred on preventing person to person transmission through social distancing or lockdown methods. The implementation of lockdowns has slowed the spread of the pandemic in many nations, to the point where the daily infection rate is well below the peak, which for many nations occurred in March/ April 2020.[4], [5] This is despite the relatively low national levels of disease immunity, according to antibody seroprevalence studies.[6]

As the first major pandemic in the information age, the global technology and connectivity that has enabled the rapid expansion of the disease, has also facilitated rapid data collection and analysis of responses to the pandemic. The international Lockdown response has been meticulously documented by the Oxford University Government Response tracker[7] which consists of individual lockdown and economic policies scored on an ordinal scale and a composite lockdown score which measures the overall lockdown strength, on a daily basis, for almost every country in the world, a sizeable undertaking which has improved our understanding of the effectiveness of Lockdown strategies.[8] While the lockdown has been widely effective, it has come with substantial financial, social and political costs and exerted a heavy toll on the mental [9], [10] and physical health [11]—[13]of the population. Perpetual lockdown is widely deemed to be politically untenable, though a lockdown of some degree may be the only way of holding the disease back and reducing mortality while vaccine development is in progress.

As many countries emerges from lockdown, there is particular concern that the course of the pandemic may echo that of the last major global pandemic, the 1919 Spanish Influenza. Over 50 million are estimated to have died, and three distinct waves were observed, the most severe being the second.[14] The conundrum of how to safely reduce or exit a lockdown policy while avoiding such a second wave is widely being evaluated and in the face of current, as far as we are able to discern, low levels of population immunity this naturally brings about anxiety and controversial policy choices.[15]

The pandemic seemed to take hold more quickly in the Northern Hemisphere, initially in Asia and European Countries in the winter and Spring months and then slowed done in the Northern hemisphere in June and July. This has coincided with acceleration of the pandemic in Southern Hemisphere winter months of June and July, at the time of writing, particularly in Brazil, Argentina, Chile, Peru, South Africa and Australia.[1] Whilst this may be due to a number of reasons, climate is likely to be factor, particularly as the transmission of respiratory pathogens such as the influenza virus is much higher in winter months when the temperature is lower. A number of studies have demonstrated that Sars-CoV-2 transmission is higher in cold weather.[16] Humidity may be an important factor, with a negative correlation to Sars-CoV-2 transmission and deaths shown in a number of studies.[17], [18]

The aim of this study is to determine the relationship between lockdown stringency and weather parameters across many nations through the use of multivariate analysis in order to generate a predictive model that could forecast future infection rates.

## Methods

The 40 countries with the highest Sars-CoV-2 cases on the 24th May 2020 was selected for the study and daily cumulative case numbers and cumulative mortality was extracted from WHO records.

National record of lockdown stringency and composite stringency index was obtained from the Oxford SARS-COV-2 Government Response Tracker. The composite is an average of 9 government responses scored on individual ordinal scales and ranges from 0 for no Lockdown to 100 for the most stringent Lockdown.

Daily temperature records were extracted from the website wunderground.org for the capital city of a nation. Data included (daily) maximum temperature, minimum temperature, and average temperature, maximum humidity, minimum humidity and average humidity, maximum dew point, minimum dew point and average dew point and precipitation for the study period (1/1/20 to 11/6/20) and for the corresponding date on the previous year for the extrapolation period of 12/6/20 to 1^st^ of July 2021. The temperature records were converted to 7 day rolling averages.

The Viral reproductive (R_0_) rate was calculated for each date by dividing the number of people who died due to Sars-CoV-2 in the following 6 days by the number of people who had died in the preceding 6 days, in keeping with infective period of 6 days and an incubation period of 6 days, similar to the method described by Heald.[4] A 7 day rolling average of R_0_ was calculated and converted to a log base 10 (LR), as this has a more normal distribution than R_0_ and prevents a negative value for R_0_ being predicted, which would be deleterious to any further extrapolation.

The relationship between weather and the viral reproductive across the 40 countries for the study period was analysed firstly using pooled data. The Pearson correlation between the 7-day rolling average weather parameter and the log10 viral reproductive rate for 0 to 35 days after the weather date, was assessed by Pearson coefficient and the optimal weather parameters and the timing of their effect on Log10 Viral reproductive rate was assessed, by maximisation of the Pearson’s coefficient. The 7-day rolling maximum temperature and 7 day rolling humidity were selected as being the most predictive.

The composite lockdown stringency index was converted to a 7-day rolling average and similarly assesses by Pearson correlation against the LR for the pooled data from 40 countries.

The study period was divided into a test period (The first 110 days from 1/1/20 to 19/04/2020) and a validation period (From 20/4/20 to 11/6/20). Multivariate linear regression was applied to data from the study test period date for 20 countries with the highest cases. Three parameters were used; 7 day rolling stringency index, 7 day rolling maximum temperature and 7-day average humidity for predictive parameters for LR. Varying time delays in the effect on LR between nations were permitted and the optimal time delay was taken as the one which was associated with the highest R^2^. For 2 countries; UK and Germany, a simpler model excluding humidity and using LR and 7 day rolling average daily temperature was used as it was found to perform better.

The multivariate equation from the test period was applied to the validation period for 11 countries with the highest case numbers. Whilst the precise coefficients were used from the test period, a change in the lag period in the determination of LR was permitted between the test and validation periods within nations. The correlation between the actual and predicted LR was determined by Pearson coefficient and the optimal was taken as that with the highest value. The calculated LR was used to calculate the rolling active infections and projected cumulative death rate and this was correlated with the actual death rate in the validation period by Pearson’s coefficient. The regression equation was used to calculate the predicted lockdown stringency required to keep R_0_ below 1 for January and July using the average whether parameters for those months.

The regression equation was used to estimate the viral reproductive rate for 11 nations between 12/6/2020 to 1/7/2021, with the predicted weather for the year deemed to being identical to the previous year. This was carried out under 4 conditions; continuation of lockdown measures present on 11/6/2020 with and without the effect of population immunity, and no lockdown with and without the effect of population of immunity. The effect of population immunity was factored by taking the fatality rate of Sars-CoV-2 infection as 0.5%, [6] and this was used to calculate the prevalence of prior infections in the community. Immunity was assumed to be homogenous across the population. The portion of the population that was susceptible was recalculated each day as ((5,000-deaths per million)/5,000)) and the effective R_0_ was taken as the actual R_0_ multiplied by the proportion of the population that was still susceptible.

With the effect of immunity, as described above, the projected cumulative death rate for 5 nations (USA, Russia, UK, Italy and India) was calculated under 1 of 4 conditions; continued current lockdown (11/6/20), 100% lockdown, 20% lockdown and No lockdown. The projected daily mortality was calculated from 12/6/20 to 1/7/21, using the formula new daily mortality (3 days later) = (prior six-day mortality) x (calculated viral rate) x (proportion of population susceptible) divided by 6. The cumulative daily re-calculations were performed on an excel spreadsheet.

All statistics were calculated using IBM SPSS. Statistical significance was taken as p<0.05 throughout.

All data is available on request from

## Results

The relationship between weather parameters were valuated against the log10 viral reproductive rate, LR, allowing for variation in the time lag to determine when the effect of weather on LR was most apparent. Pooled data for 40 countries from 1/1/20 to 24/5/20 was utilised. Maximum daily temperature and average humidity had the highest correlation with LR (data not shown). Figure 1 shows the relationship between 7 day rolling maximum temperature and humidity and precipitation and LR. Maximum temperature had a negative correlation with LR. Peak Pearson correlation =-0.174 (n=2907, p=0.000) at 2 days later however, this is too soon to be due to viral transmission. A second mini peak occurs 14 days later, Pearson coefficient =-0.140 (n=2907, p=0.000). Precipitation was associated with viral transmission with a peak effect 20 days later, coefficient = 0.098 (n=2906, p=0.000). Humidity had the strongest effect on this univariate association with a first peak at 0.233 at 21 day later (n=2906, p=0.000 and a higher second peak of association at 33 days later 0.263 (n=2903, p=0.000).

**Figure 1:**
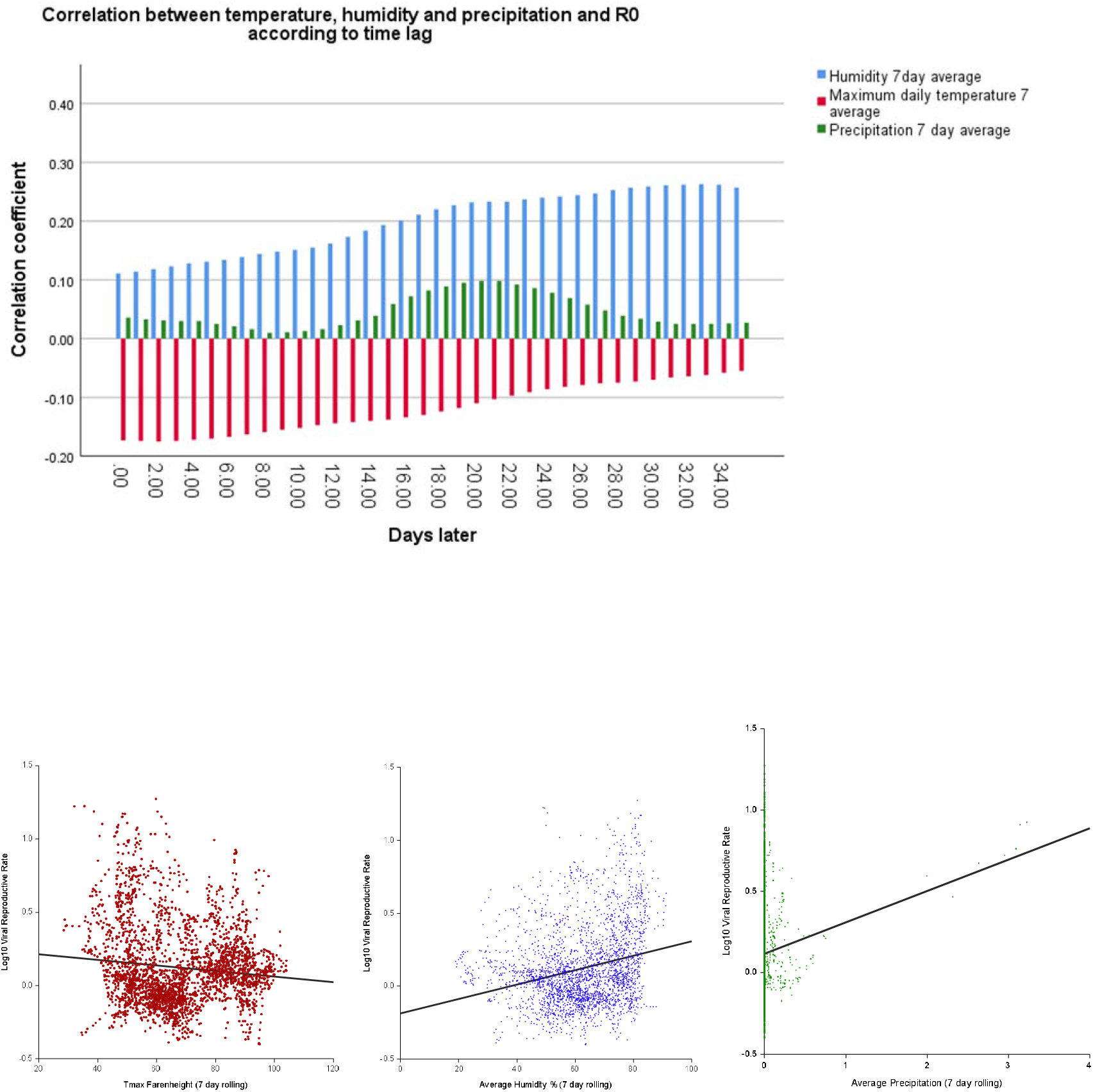
The relationship between Temperature, humidity, and precipitation and log_10_ viral reproductive rate, from 40 countries 1/1/20 to 11/6/20 (n=2907). A) Change in Pearon’s correlation coeffcient with time lag after weather parameter. B-D; Scatter plots of 7 day rolling mean of daily maximum temperature (Fahrenheight, r^2^=0.01); 7 day day rolling mean humidity (%, r^2^ =0.054); 7 day rolling preciptation (mm, r^2^=0.01) and log_10_ viral reproductive rate 21 days later.

**Figure 2:**
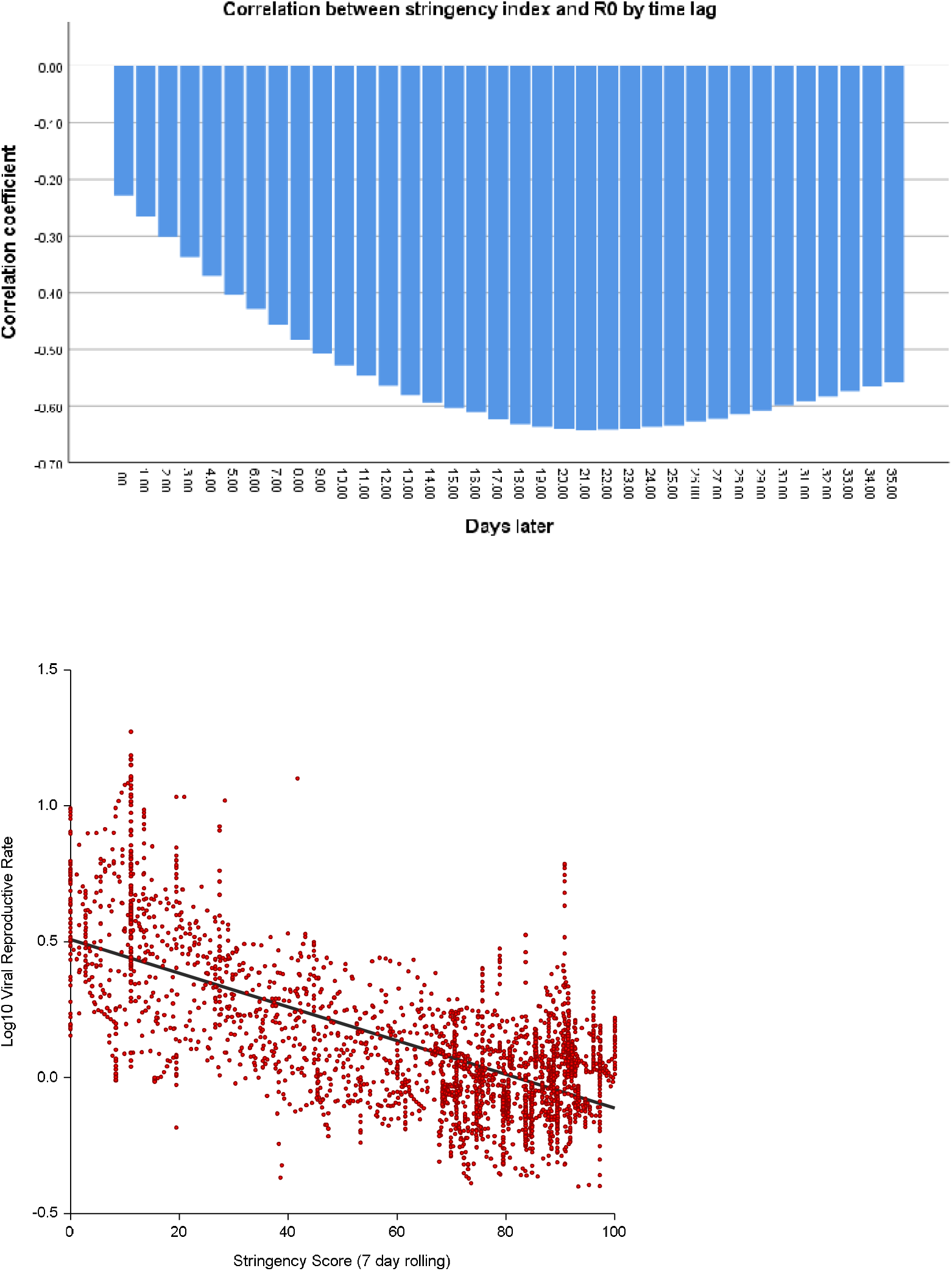
T The relationship between rolling 7 daylockdown stringency and log_10_ viral reproductive rate, from 40 countries 1/1/20 to 11/6/20 (n=2907). A: change in Pearson’s coefficient according to time lag and B) Scatter plot of rolling 7 day lockdown stringency versus log_10_ viral reproductive 22 days later (n=2906, r^2^=0.42).

The relationship between stringency index and LR was evaluated from the poled date for 4-nations. The rolling 7-day stringency index has a much more strongly predictive of LR than the weather parameter with a strong negative correlation. Pearson correlation = -0.641 peaking at 22 days (n=2898, p=0.000).

Multivariate regression for LR for the test period was carried out for the 20 countries with the highest Sars-CoV-2 test numbers on the 24/5/2020. Table 1 tabulates the calculated coefficients for the regression equation using rolling 7-day stringency index, rolling 7-day average of maximum temperature and rolling 7-day average of humidity. A second model was created for the UK and Germany using only stringency and average humidity and was used in subsequent analyses. The stringency index had a negative correlation with LR in all countries, and was statistically significant (p<0.05) in 19 out of 20 countries evaluated. Maximum temperature had a negative correlation in 17 out of the 20 countries and was statistically significant in 16 out of 20 countries. In these multivariate analyses, humidity faired relatively poorly when compared to the univariate correlation achieved for pooled data. It was positively associated in 11 out of 20 countries and significantly associated in 13 out of the 20 countries. The R^2^ for the test model varies from 0.564 for China to 0.987 for Italy and averages 0.889. The time lag for the peek predictability of the equation varied from 9 to 32 days.

**Table 1.** Summary of coefficient used in multivariate regression prediction of log_10_viral reproductive rated for 20 countries with highest incidence of Sars-CoV-2 at time of evaluation, for the model generation period (1/1/20 to 19/4/20) with respective time delays of prediction. * Average temperature used.

| | Coefficients of fitted equation for $\log_{10} R_0$ | | | | | | Time Lag |
| --- | --- | --- | --- | --- | --- | --- | --- |
| Country | Constant<br>+95% CI | Stringency Index<br>+95% CI | Maximum<br>temperature<br>+95% CI | Average Humidity<br>+95% CI | R<sup>2</sup> for R<sub>0</sub>, SE of<br>regression<br>estimate | N | Days |
| USA | 1.649<br>(1.229, 2.069) | -0.007<br>(-0.009, -0.005) | -0.013<br>(-0.019, -0.007) | -0.006<br>(-0.008, -0.004) | 0.903, 0.094 | 73 | 27 |
| Russia | 1.325<br>(1.07, 1.60) | -0.006<br>(-0.006, -0.006) | -0.012<br>(-0.006, -0.018) | -0.003<br>(-0.001, -0.005) | 0.856, 0.061 | 47 | 27 |
| Brazil | -1.05<br>(-2.48, 0.38) | -0.012<br>(-0.014, -0.010) | 0.014<br>(0.004, 0.024) | 0.013<br>(0.001, 0.025) | 0.854, 0.072 | 38 | 9 |
| Spain | 0.399<br>(-0.315, 1.113) | -0.016<br>(-0.018, -0.014) | 0.002, ns<br>(-0.006, 0.010) | 0.012<br>(0.004, 0.020) | 0.938, 0.11 | 51 | 9 |
| UK | 2.161<br>(1.823, 2.499) | -0.010<br>(-0.010, -0.010) | -0.026*<br>(-0.034, -0.018) | Not utilised | 0.923, 0.093 | 47 | 7 |
| Italy | 1.326<br>(1.120, 1.532) | -0.012<br>(-0.012, -0.012) | -0.006<br>(-0.008, -0.002) | 0.001, ns<br>(-0.001, 0.002) | 0.987, 0.037 | 70 | 17 |
| France | 1.488<br>(1.08, 1.88) | -0.014<br>(-0.015, -0.013) | 0.002, ns<br>(-0.002, -0.006) | -0.009<br>(-0.013, -0.005) | 0.946, 0.092 | 70 | 21 |
| Germany | 1.703<br>(1.59, 1.80) | -0.014<br>(-0.015, -0.014) | -0.014*<br>(-0.017, -0.011) | Not utilised | 0.986, 0.039 | 52 | 14 |
| Turkey | 1.737<br>(1.141, 2.333) | -0.013<br>(-0.015, -0.011) | -0.013<br>(-0.019, -0.007) | 0, ns<br>(-0.004, 0.004) | 0.917, 0.080 | 43 | 15 |
| Iran | 5.694<br>(4.802, 6.547) | -0.008<br>(-0.002, -0.014) | -0.049<br>(-0.059, -0.039) | -0.033<br>(-0.039, -0.027) | 0.926, 0.074 | 39 | 11 |
| India | 2.415<br>(1.33, 3.50) | -0.006<br>(-0.008, -0.004) | -0.012<br>(-0.020, -0.004) | -0.011<br>(-0.017, -0.005) | 0.817, 0.086 | 57 | 23 |
| Peru | 3.655<br>(0.71, 6.59) | -0.004<br>(-0.004, -0.004) | -0.04<br>(-0.062, -0.016) | 0.003, ns<br>(-0.011, 0.017) | 0.823, 0.075 | 56 | 29 |
| China | 4.838<br>(3.53, 6.13) | -0.002, ns<br>(-0.008, 0.004) | -0.06<br>(-0.076, -0.044) | -0.032<br>(-0.056, -0.018) | 0.564, 0.46 | 92 | 3 |
| Canada | 0.268<br>(0.04, 0.48) | -0.005<br>(-0.005, -0.005) | -0.008<br>(-0.006, -0.010) | 0.006<br>(0.004, 0.010) | 0.935, 0.06 | 61 | 31 |
| Saudi Arabia | 3.353<br>(2.89, 3.81) | -0.026<br>(-0.030, -0.022) | -0.008<br>(-0.014, -0.002) | -0.005<br>(-0.007, -0.003) | 0.919, 0.054 | 32 | 10 |
| Mexico | 6.126<br>(4.89, 7.40) | -0.002<br>(-0.002, -0.002) | -0.061<br>(-0.075, -0.047) | -0.021<br>(-0.027, -0.051) | 0.870, 0.068 | 51 | 25 |
| Chile | -0.333<br>(-1.59, 0.97) | -0.006<br>(-0.006, -0.006) | 0.008, ns<br>(-0.004, 0.020) | 0.004, ns<br>(-0.002, 0.010) | 0.946, 0.043 | 43 | 19 |
| Belgium | 2.87<br>(2.17, 3.58) | -0.014,<br>(-0.016, -0.012) | -0.024<br>(-0.020, -0.028) | -0.004, ns<br>(-0.010, 0.002) | 0.954, 0.098 | 45 | 10 |
| Pakistan | 1.992<br>(0.49, 3.49) | -0.004<br>(-0.006, -0.002) | -0.01, ns<br>(-0.022, 0.002) | -0.009, ns<br>(-0.019, 0.001) | 0.752, 0.055 | 37 | 10 |
| Netherlands | 0.653<br>(0.49, 0.81) | -0.009<br>(-0.011, -0.007) | -0.004<br>(-0.006, 0) | 0.005<br>(0.001, 0.009) | 0.971, 0.064 | 52 | 12 |

Evaluation of regression model of LR for validation period; the equation from table 1 was applied to the validation period 20/4/20 to 11/6/20 for the 11 countries with the highest case numbers. Correlations between the equation and differing time delays for LR were permitted and the optimal correlation is shown in Table 2. On average, the time lag for peak prediction changed from 16.3 to 19.5 days between the test and validation period. The resultant Pearson coefficient for LR for the validation period ranges from 0.344 for Germany to 0.919 for Italy. The predicted model is shown versus the actual viral reproductive rate as time series for 5 nations in Figure 3.

**Table 2.** Summary of model (from table 1 but with varied time lag) with predictivity for validation period (19/4/20 to 11/6/20) of viral reproductive rate and cumulative mortality.† Spanish validation period form 19/4/20 to 13/5/20 due to downward revision of national mortality data on the 24/5/20. Predicted lockdown stringency to keep R_0_ below 1 for January and July shown (excluding immunity). Predicted cumulative mortality for 1/7/21 with 11/6/20 lockdown stringency perpetuated from 11/6/20 onwards.

| Country | Pearson correlation for $\log_{10} R_0$ (19/4/20 to 11/6/20), (95%CI) | n | Time lag validation period | Pearson correlation for Total deaths from 1/1/20 to 11/6/20, 95% CI | Required lockdown stringency July | Required lockdown stringency January | Projected Total mortality on 1/7/21 with 11/6/20 lockdown continued |
| --- | --- | --- | --- | --- | --- | --- | --- |
| USA | 0.786<br>(0.571, 0.894) | 27 | 20 | 1.000<br>(1.000, 1000) | 12.0 | 87.6 | 117865 |
| Russia | 0.834<br>(0.634, 0.924) | 23 | 27 | 0.999<br>(0.999, 0.999) | 45.2 | 100+ | 380268 |
| Brazil | 0.743<br>(0.534, 0.860) | 34 | 16 | 0.982<br>(0.975, 0.986) | 90.5 | 97.9 | 608373 |
| Spain | 0.730†<br>(0.454, 0.870) | 24 | 8 | 0.999<br>(0.999, 0.999) | 61.0 | 91.0 | 30373 |
| UK | 0.653<br>(0.399, 0.807) | 34 | 16 | 0.999<br>(0.999, 0.999) | 37.0 | 93.9 | 45053 |
| Italy | 0.919<br>(0.817, 0.962) | 25 | 25 | 1.000<br>(1.000, 1.000) | 70.6 | 88.8 | 193692 |
| France | 0.703<br>(0.463, 0.840) | 32 | 18 | 1.000<br>(1.000, 1.000) | 84.0 | 57.5 | 30258 |
| Germany | 0.344<br>(0.035, 0.587) | 40 | 11 | 1.000<br>(1.000, 1.000) | 53.5 | 81.4 | 9325 |
| Turkey | 0.891<br>(0.695, 0.958) | 18 | 34 | 0.980<br>(0.972, 0.985) | 48.8 | 85.0 | 9446 |
| Iran | 0.529<br>(0.239, 0.725) | 36 | 14 | 0.999<br>(0.999, 0.999) | 0 | 100+ | 419375 |
| India | 0.619<br>(0.355, 0.784) | 35 | 15 | 0.991<br>(0.988, 0.994) | 77.0 | 100+ | 5020424 |

**Figure 3:**
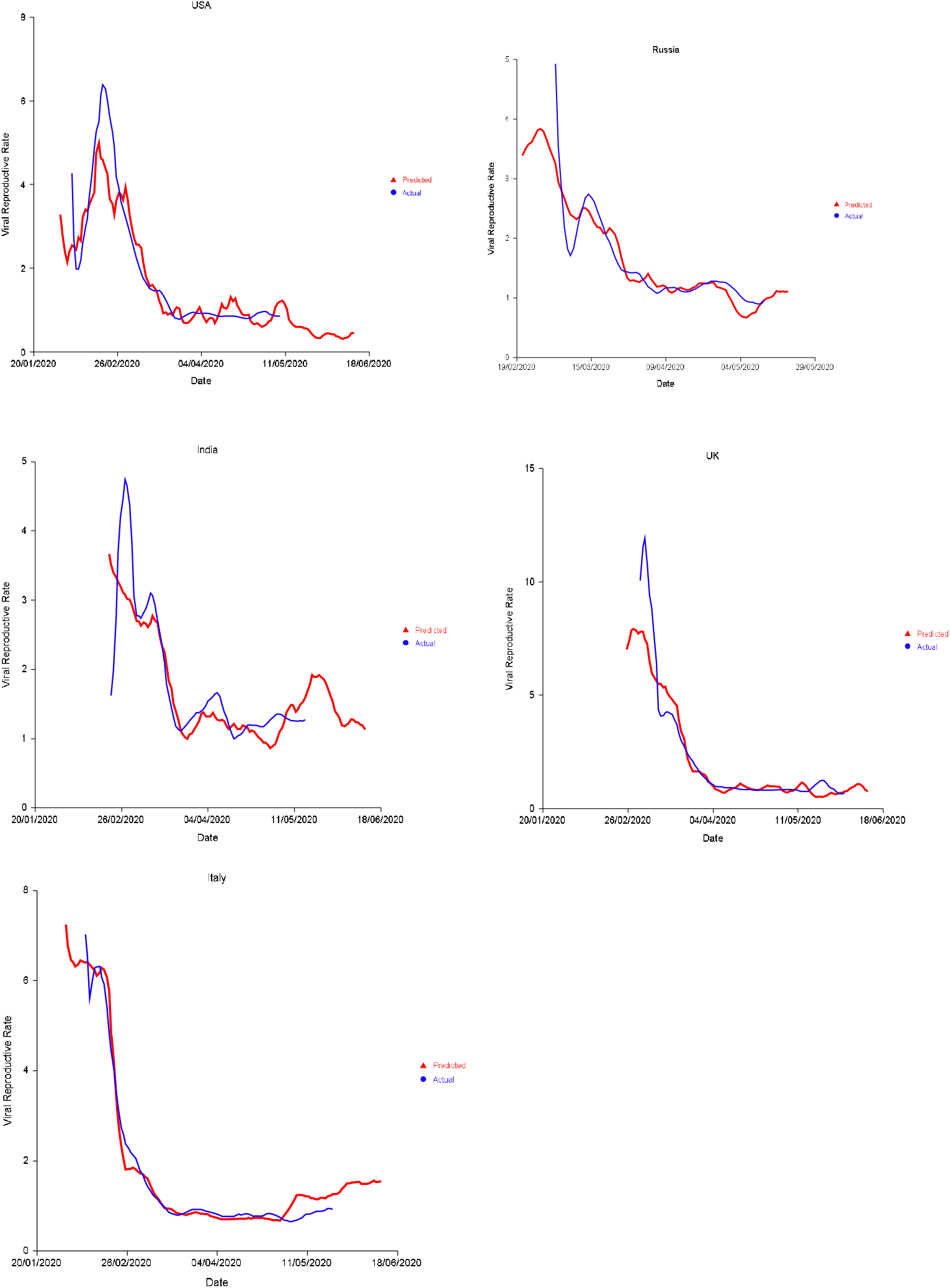
Calculated viral reproductive rate versus actual rate for selected countries. Model generated 1/1/20 to 19/4/20 and extrapolated 20/4/20 to 11/6/20 (Lag time for test period used throughout test and validation period in graphs below).

The daily predicted viral reproductive rate was used to calculated cumulative predictive mortality for each country. This show a very strong correlation with mortality for the test period with Pearson coefficient ranging 0.980 for Turkey to 1.000 for Italy, France Germany and USA, and is shown graphically in Figure 4 for 5 selected countries as a time series and as a correlation between predicted and actual mortality.

**Figure 4.**
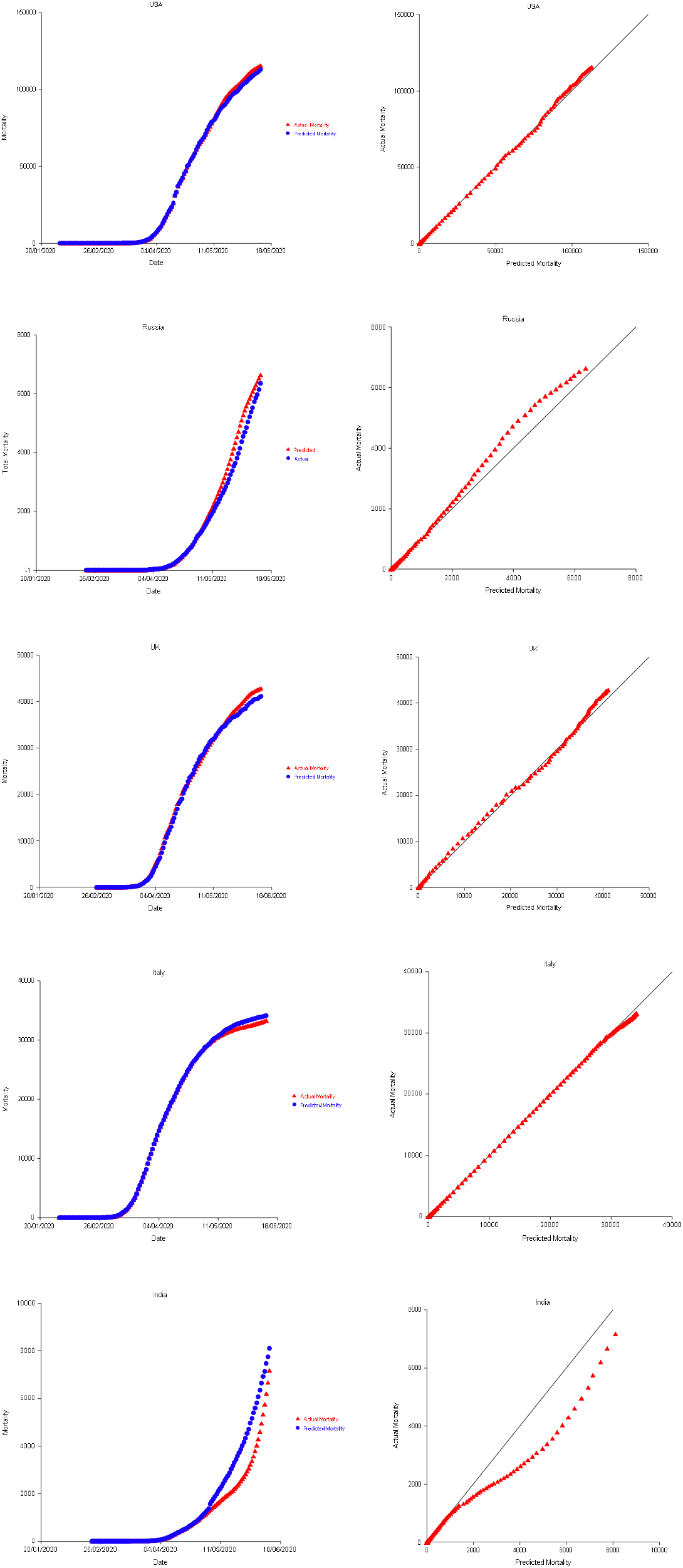
Calculated cumulative mortality versus actual mortality for selected countries. Extrapolation period 20/4/20 to 11/6/20. 45 degree line represents perfect correlation.

The predicted lockdown stringency score to keep R_0_ at 1 or less is shown for the 11 nations in table 2 for January and July. The prediction assumes 100% population susceptibility. The average lockdown stringency required in January is 95.8, and the average lockdown stringency required in July is 51.0. All countries apart from France are predicted to require a stronger lock down in January than July. USA and Iran are predicted to require minimal lockdowns during July.

Long term (from 12/6/20 to 1/7/2021) prediction of viral reproductive rate and cumulative mortality is shown in Figure 5 for 5 nations. Viral reproductive rate is shown under 4 conditions; current lockdown without immunity factored, current lockdown with immunity factored, no lockdown without immunity factored and with no lockdown with immunity factored. Cumulative mortality (with immunity factored) is shown under 4 conditions; current lockdown applied throughout (from 11/6/20 onwards), 100% lockdown applied throughout, 20% lockdown applied throughout and no lockdown applied throughout. In the case of UK and USA, the current lockdown (on 11/6/20), if perpetuated, is sufficient to almost completely curtail the spread of the virus. The model predicts that with current lockdown, Italy and India would experience resurgence of viral transmission towards the end of 2020 which could be abolished with a more stringent lockdown. In Russia, with the current lockdown resurgence is delayed to February 2021. In all cases no lockdown or 20% lockdown results in spread to 80-90% of the population (taking the fatality rate as 0.5%) within 2020 or the start of 2021. The final cumulative mortality estimate for July 1^st^ 2021 with current (11/6/20) lockdown perpetuated and population immunity factored is tabulated in the final column of table 2. This may be less than more current mortality data as national lockdowns have been subsequently eased from the time of the analysis.

**Figure 5;.**
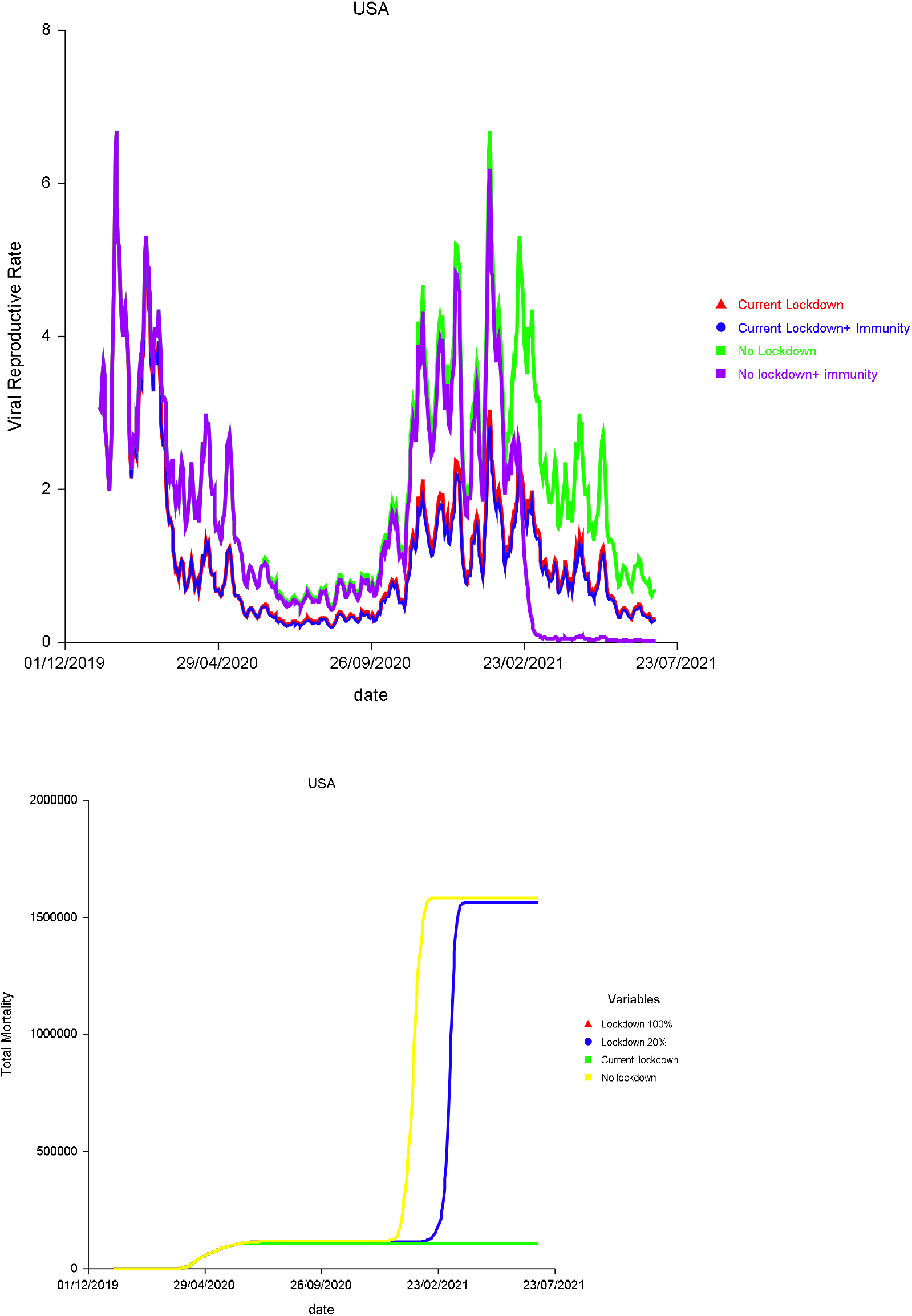
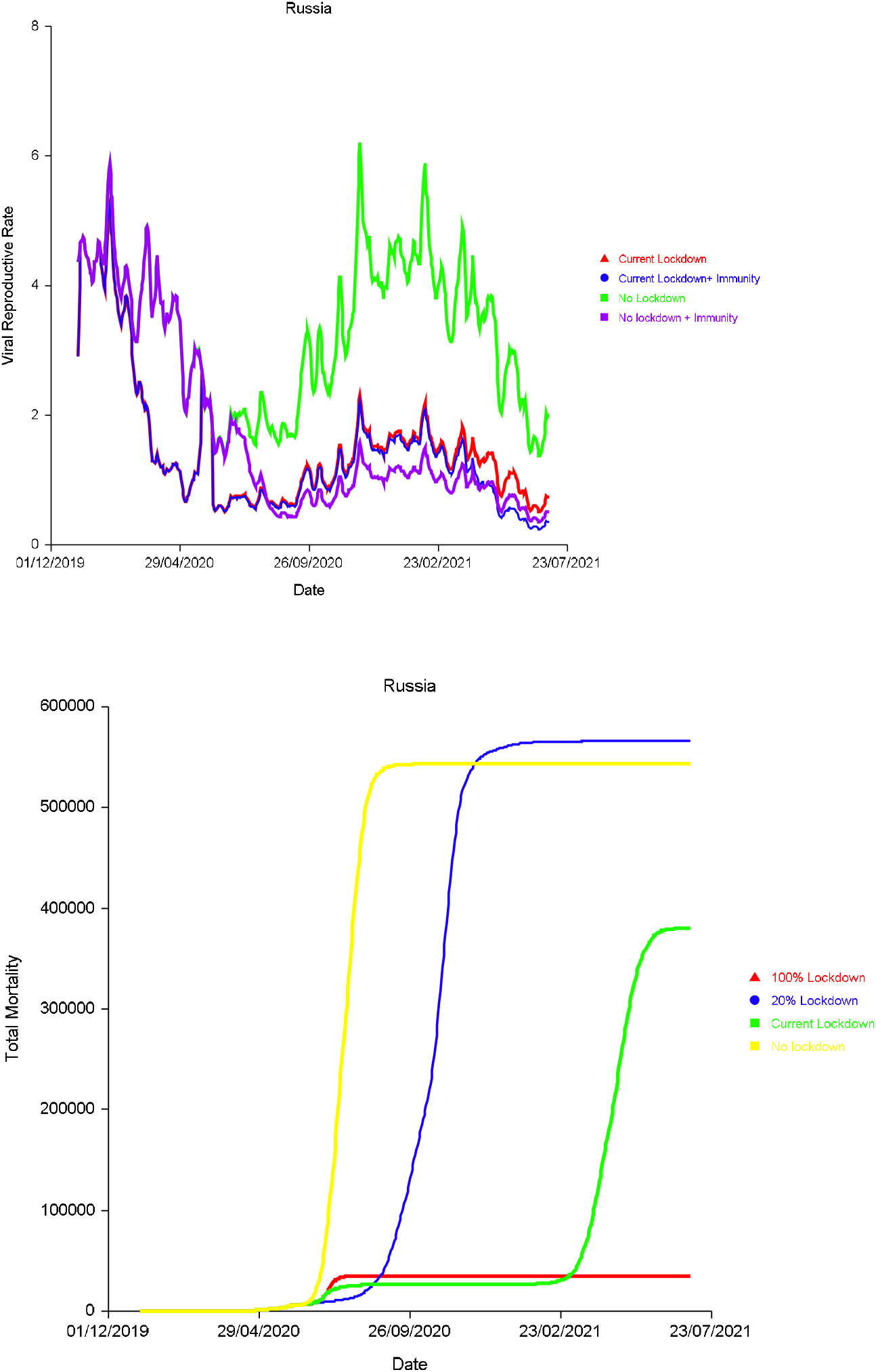
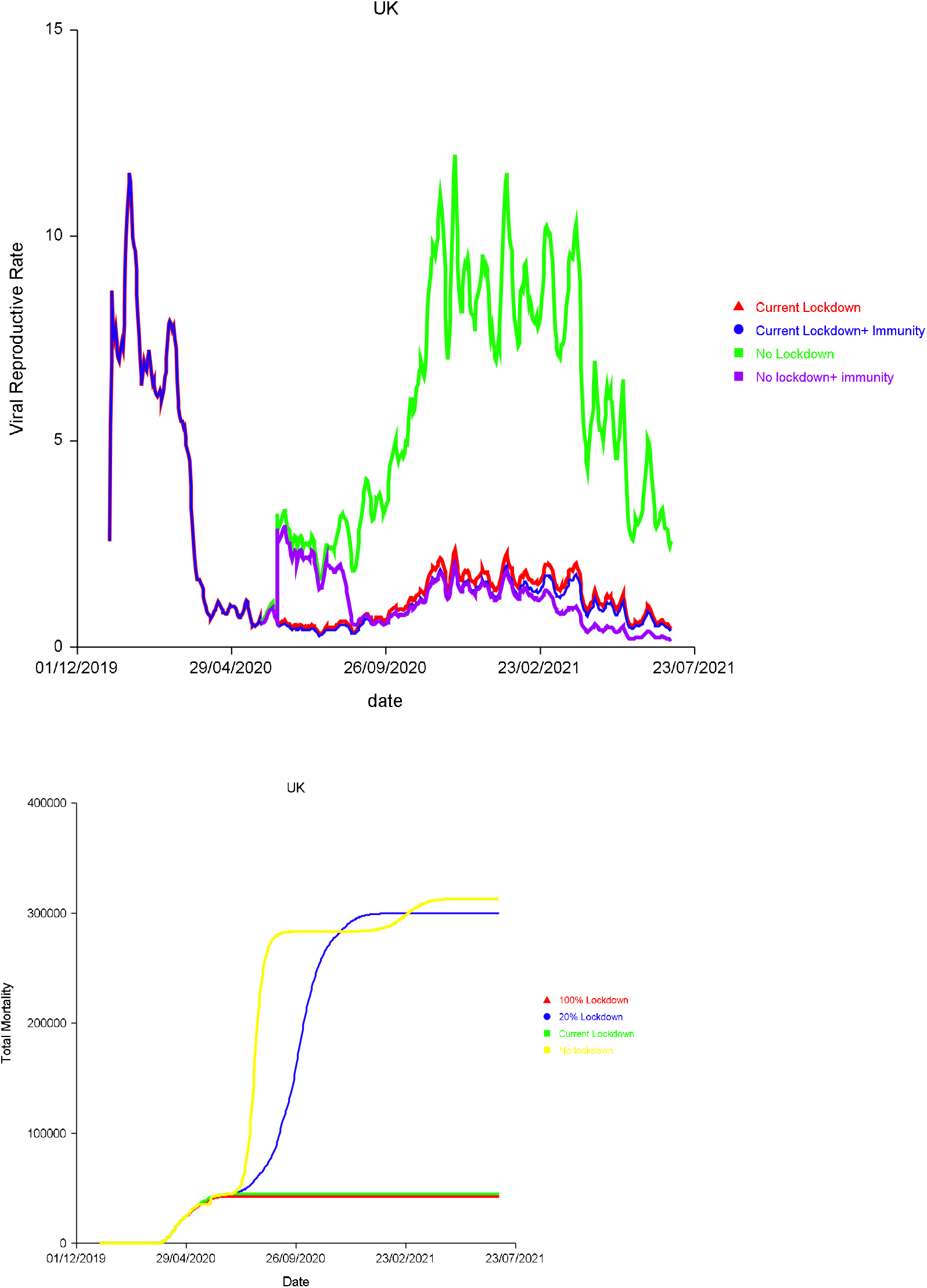
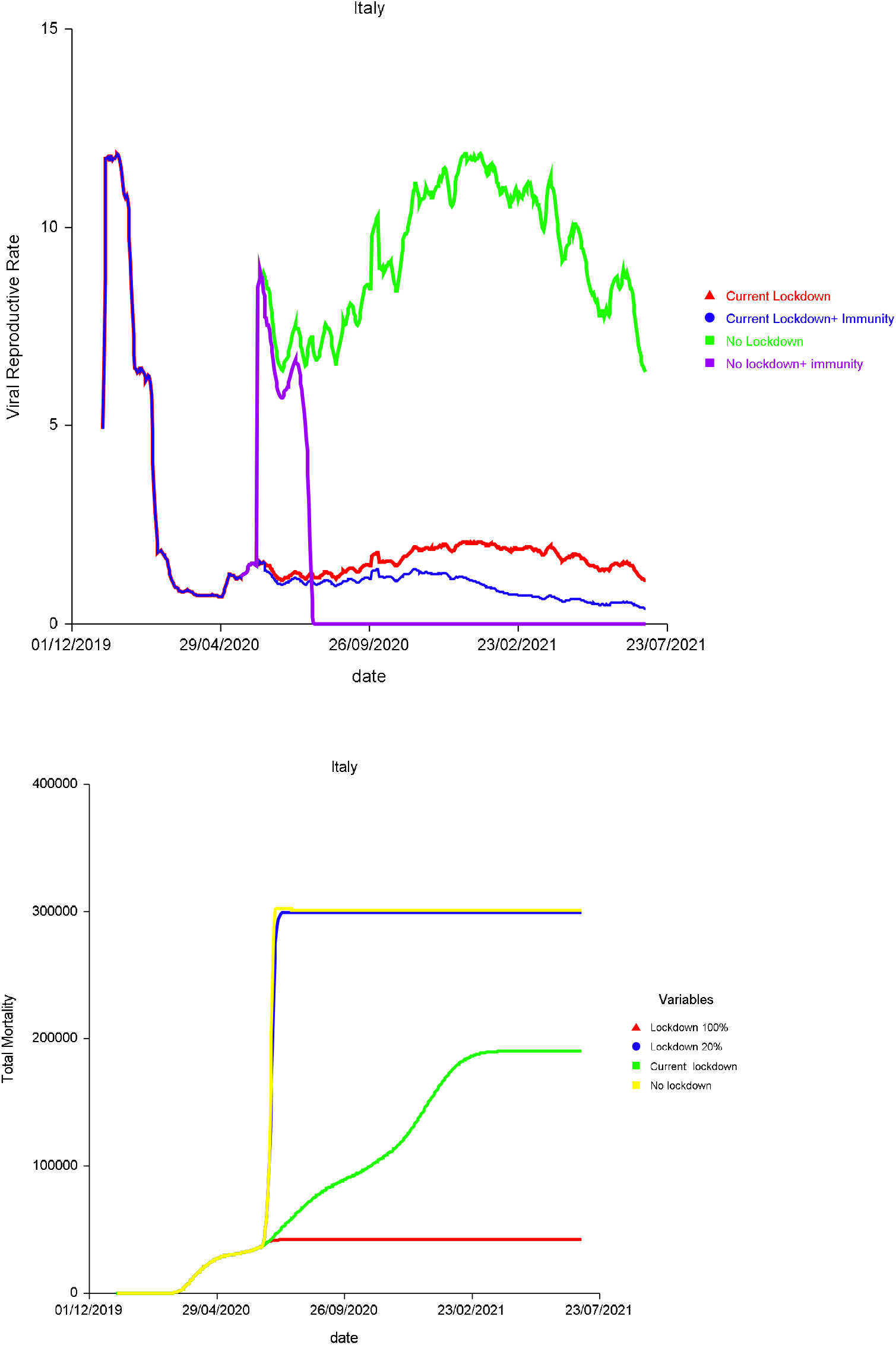
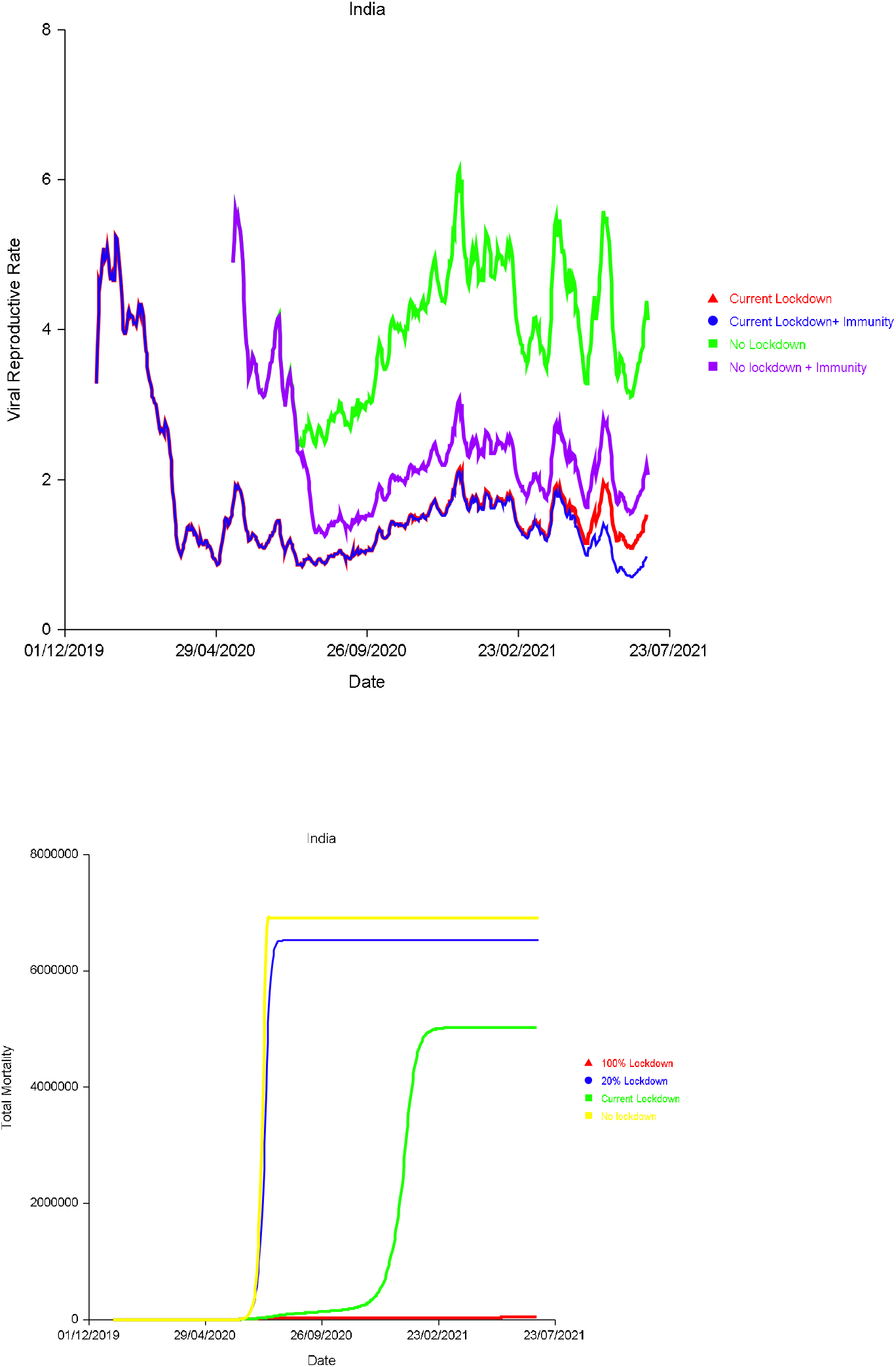
Model extrapolation for 5 selected countries, demonstrating; A) predicted viral reproductive rate for 1) current lockdown (11/6/20), 2) current lockdown with correction for immunity 3) No lockdown 4) No lockdown corrected for immunity and: B) Predicted mortality (immunity corrected in all cases) for 1) 100% lockdown, 2) 20% lockdown, 3) current lockdown (11/6/20) and 4) No lockdown.

## Discussion

This is the first predictive model which assesses the effect of lockdown and weather parameters on the Sars-CoV-2 viral reproductive rate, demonstrating the size of their effects and the time delay which determines these relationships.

Of the three parameters evaluated, the lockdown was most predictive of the viral reproductive rate. From the pooled date the peak association was -0.641 at 22 days. In the individual country analysis from the 20 countries with the most cases (Table 1), lockdown stringency score was a significant predictor in 19 countries.

Increased humidity was strongly associated with increased viral transmission in the pooled data with a later peak of 0.263 association at 33 days, perhaps reflecting fomite transmission which could be relatively delayed. However, in the individual country multivariate analysis, humidity did not perform so well as a predictive factor. It was omitted from the Germany and UK model where the average daily rolling temperature was used. Humidity had a positive relationship with viral transmission in some countries and a negative relationship in others, therefore the relationship with humidity is not clear. The difference between the pooled and individual analysis could be due to; the relationship between the average humidity and some other confounding variable which differs between countries; its relationship which temperature which may supervene on multivariate analysis; this study not being optimised to examine the effect of humidity, as for each country a single overall time lag was chosen which averaged 16.5 days in the validation period, and away from the peak of association of humidity, which was found to be at 33 days. Some studies show a bell-shaped curve conferring lower virus survival at moderate levels of humidity,[19] potentially explained by the behaviour of salts dissolved within the respiratory droplets.[20] Depending on the range of humidity experienced by a certain country, this may not be apparent in our model.

Temperature had a negative association from pooled data and was typically negative in the individual country multivariate analysis. This aligns with other studies of transmission, and is to be expected due to the widely documented inactivation of coronaviridae at higher temperatures[21], [22] and reduced lifespan of respiratory droplets.[23]

As it is widely believed that previous Sars-CoV-2 infection will confer immunity against re-infection during the pandemic, the cumulative incidence of disease, has an important bearing on any forecast model. While questions remain regarding the longevity of such an effect,[24] even a small degree of immunity could reduce the effective viral rate beneath 1, in which case further disease spread would be limited. Calculations using the viral reproductive rate are compound in nature, and a small variation in the number of infections at one point in time will have a large bearing on the number of infections at a later date. However, we believe any predictive model should centre on prediction of the viral reproductive rate, which though widely variable, is predicted by the variables evaluated here, in particular the degree of lockdown. As the viral reproductive rate fluctuates form day to day, it is difficult to predict, nonetheless the predicted equations for the test period had a typically high R^2^ (up to 0.987 with standard error of 0.037) for predicting the log of the viral reproductive, with equates to a 95% Confidence interval of 0.84 to 1.18 for a predicted viral rate of 1. When applied to the extrapolation period there is high correspondence with the predicted viral rate ranging from 0.344 to 0.919, all be it with a change in the time delay for peak correlation between lock down and temperature parameters and the viral reproductive rate. When this predicted viral rate is used to calculate the predicted death rate for the validation period, the strength of association is greater than the viral reproductive rate despite this being a compound calculation. For the validation period, in most countries, the viral reproductive rate was close to 1 and the log_10_ was close to zero so on a logarithmic scale, the size of errors can seem magnified in relation to the size of the value predicted. For long term predictions over many months, the change in time lag between the validation and test period makes little difference. A change in the time lag between the effect of lockdown and weather on the observed viral reproductive rate, could be due to change in reporting practices over time and survivability as the R_0_ was estimated using the mortality numbers rather than case numbers.

The extrapolation models for the selected countries predict that reducing the lockdown stringency to 20% or less would be insufficient to control the pandemic and this accords with previous findings, where it had been predicted that most countries would need to spend > 80% of time in their respective lockdown to prevent the pandemic from resurging.[5] In the countries evaluated by our model, in the absence of social distancing, the pandemic could resurge at any time, but there is a greater propensity for this in winter months and a more stringent lockdown would be required for these periods. The model predicts that USA, and Iran would require minimal lockdown in the July whilst Brazil France and India would still require strong lockdowns in July. All the countries evaluated, aside from France, required stronger lockdowns in January than July. According to the model, in India, Russia and Brazil, much stronger lockdowns than previously employed are required. The measures taken to combat the pandemic need to be seen as part of a long term strategy and the findings of this model could facilitate long term planning, whilst vaccine evaluations are in progress.

The predicted mortality curves are theoretical and unlikely to representative the actual long term course of the pandemic. This is due to the effect of cumulative errors over time, changes in government policies and behaviour in response to viral transmission and the case fatality rate, which is likely to decrease in response to improved treatment and novel therapies, such as the use of Dexamethasone emerging from the RECOVERY trial.[25] There is evidence that the ITU mortality rate has decreased over time,[26] and new trialled therapies such as nebulised Interferon Beta may confer significant survival benefits.[27] Even in the absence of governmental instructions, in the case of a resurgence of the disease, a large proportion of individuals are likely to implement their own measures such as avoiding busy locations and enhanced hygiene practices, akin to a degree of lockdown.[28]

Weaknesses of this study include use of temperature from the capital city which may vary widely across regions, the relatively small number of data points for multivariate analysis, the limitations of recorded Sars-CoV-2-19 data, and the heterogeneity between interpretation and application of lockdown policies between countries.

### In conclusion

The viral reproductive rate for Sars-CoV-2-19 can be accurately predicted using a measure of Lockdown stringency and weather parameters.

## Data Availability

All data is available upon request from

